# Early death after a diagnosis of metastatic solid cancer – raising awareness and identifying risk factors from the SEER database

**DOI:** 10.1101/2023.01.27.23285089

**Authors:** Opher Globus, Shira Sagie, Noy Lavine, Daniel Itshak Barchana, Damien Urban

**Affiliations:** Institute of Oncology, Sheba Medical Center, Ramat Gan, 52621, Israel; Sackler Faculty of Medicine, Tel Aviv University, Tel Aviv, Israel; The Sheba Talpiot Medical Leadership Program; Department of Molecular Cell Biology, Weizmann Institute of Science, 7610001 Rehovot Israel; St. George’s University of London medical program delivered by University of Nicosia Medical School, Nicosia, Cyprus

**Keywords:** metastatic cancer, solid tumors, early mortality

## Abstract

**Background:** Cancer death rates are declining, in part due to smoking cessation, better detection and new treatments; nevertheless, a large fraction of metastatic cancer patients die soon after diagnosis. Few studies and interventions focus on these patients. Our study aims characterize early mortality in a wide range of metastatic solid tumors.

**Methods:** We retrieved data on adult patients diagnosed with pathologically confirmed *de-novo* metastatic solid tumors between the years 2004-2016 from the Surveillance, Epidemiology, and End Results database (SEER). Our primary outcome was cancer specific early death rate (defined as death within two months of diagnosis). Additional data extracted included socio-demographical data, tumor primary, sites of metastases, and cause of death.

**Results:** 109,207 (20.8%) died of cancer within two months of diagnosis. The highest rates of early death were found in hepatic (36%), pancreato-biliary (31%) and lung (25%) primaries. Factors associated with early death included primary site, liver and brain metastases, increasing age and lower income. Cancer was the cause of death in 92.1% of all early deaths. Two-month mortality rates have improved during the study period (from 22.4% in 2004 to 18.8% in 2016).

**Conclusion:** A fifth of metastatic cancer patients die soon after diagnosis, with little improvement over the last decade. Further research is required to better classify and identify patients at risk for early mortality, which patients might benefit from faster diagnostic tracks, and which might avoid invasive and futile diagnostic procedures.

## Introduction

Despite improvements in early detection and treatment, cancer is the second leading cause of death in the United States (1). Major advances in the personalization of systemic treatment of cancer has demonstrated rapid responses and long-term survival in some patients, even in the metastatic setting (2). However, clinicians, often encounter patients with suspected *de-novo* metastatic disease who are extremely ill at the time of presentation. In these clinical situations, physicians, patients, and families are faced with fundamental decisions. On the one hand, particularly due to advances in personalized oncology, some select patients can rapidly improve with targeted treatments. On the other hand, diagnostic procedures are not without financial costs and potential complications (3), often resulting in hospital admissions and poor end-of-life care. As many of the pivotal improvements require predictive biomarkers, which can take weeks to return, the benefit of these treatment advances may elude many patients.

The aim of our current study is to describe early mortality in solid tumors and to characterize which patients with *de-novo* metastatic disease die within two months of diagnosis. As early mortality from cancer is due to a combination of complex tumor related, patient related, and health-care related mechanisms, our objective is to present the scope of this phenomenon to health care professionals involved in the diagnosis and treatment of cancer and to identify characteristics associated with early mortality, thereby helping guide physicians, patients, and families in appropriate clinical decisions.

## Methods

### Study population and design

Patients with histologically confirmed *de novo* metastatic cancer were identified from the Surveillance, Epidemiology, and End Results (SEER) database of the National Cancer Institute from 2004 to 2016. The 18 population based SEER cancer registries cover 28% of the US population (4). Given the nature of this study, there was no requirement for institutional review board submission. Access to the SEER data was in accordance with the SEER data agreement.

Patients included were at least 18 years of age and presented with a first diagnosis of pathologically confirmed metastatic cancer. Patient with recurrent metastatic disease were excluded. As the focus of our study was metastatic solid tumors, we excluded patients with primary neurological tumors, hematological malignancies, and rare solid tumors (peripheral nervous system tumors, tumors stemming from the retroperitoneum, non-lung cancer chest tumors, tumors identified as not otherwise specified from the male genital, female genital, and the urinary tract).

### Study variables

Our primary outcome was cancer specific two-month mortality from the date of diagnosis. Additional data extracted included age, sex, race, residence, income, tumor primary site, metastasis location (available only after 2010 for liver, bone, brain, and lung), year of diagnosis (grouped in four time periods 2004-2006, 2007-2009, 2010-2012, 2013-2016), cause of death, and survival by months.

We separately analyzed twelve groups of tumor locations: bladder, breast, colorectal, gastro-esophageal, hepatocellular, head and neck, kidney, lung, melanoma, ovary, pancreato-biliary and prostate, all defined by primary tumor location except for melanoma which was defined by both histology and location.

### Statistical analysis

The Mann-Whitney *U* test was used for variables that did not follow a normal distribution and the chi square test and Fisher Exact test were used as appropriate for categorical variables. We fitted multivariable logistic models. Data for metastasis sites was available since 2010, thus we fitted separate multivariable models for the whole population and for patients diagnosed after 2010. Adjusted OR and 95% confidence intervals (CI) were obtained for the independent variables from these models. P<0.05 was considered statistically significant. The statistical analyses were performed using R version 4.0.3.

## Results

### Baseline characteristics

We identified 525,780 patients diagnosed with de-novo metastatic solid malignancies between 2004 and 2016 of which 109,207 (20.77%) died of cancer within two months of diagnosis. Patient characteristics are presented in table 1. Most patients were male (52.1%), white (78.6%), and 50-70 years old (51.7%). The most common primary site was lung (40%) followed by pancreato-biliary (11.2%) and colorectal cancer (10.8%). Patients that died early were older (70 vs 65), more frequently male (54.3% vs 51.5%), had lower income (56.8% vs 54.9% had an income below $65K a year) and more frequently had brain (15% vs 12.9%), lung (49.2% vs 33.5%) and liver metastasis (33.5% vs 28%).

**Table 1:** Characteristics of the study group classified by early mortality

|  |  | **Early mortality** |  |  |
| --- | --- | --- | --- | --- |
|  | **Overall** | **No** | **Yes** | **p** |
| N | 525780 | 416573 | 109207 |  |
| Sex, Women (%) | 273976 (52.1) | 214661 (51.5) | 59315 (54.3) | <0.001 |
| Race (%) |  |  |  | <0.001 |
| White | 413207 (78.6) | 325598 (78.2) | 87609 (80.2) |  |
| Black | 69371 (13.2) | 55525 (13.3) | 13846 (12.7) |  |
| American Indian/Alaska Native | 3576 (0.7) | 2814 (0.7) | 762 (0.7) |  |
| Asian or Pacific Islander | 38627 (7.3) | 31769 (7.6) | 6858 (6.3) |  |
| Unknown | 999 (0.2) | 867 (0.2) | 132 (0.1) |  |
| Age (Median [IQR])) | 66 [57, 75] | 65 [56, 74] | 70 [61, 78] | <0.001 |
| Residence Category (%) |  |  |  | <0.001 |
| Metropolitan | 313730 (59.8) | 248942 (59.8) | 64788 (59.4) |  |
| Rural | 27729 (5.3) | 21788 (5.2) | 5941 (5.4) |  |
| Urban | 183506 (35.0) | 145222 (34.9) | 38284 (35.1) |  |
| Income below \$65K/year (%) | 284945 (55.3) | 224293 (54.9) | 60652 (56.8) | <0.001 |
| Grouped Years (%) |  |  |  | <0.001 |
| 2004-2006 | 113549 (21.6) | 88288 (21.2) | 25261 (23.1) |  |
| 2007-2009 | 118303 (22.5) | 93352 (22.4) | 24951 (22.8) |  |
| 2010-2012 | 119990 (22.8) | 95143 (22.8) | 24847 (22.8) |  |
| 2013-2016 | 173938 (33.1) | 139790 (33.6) | 34148 (31.3) |  |
| Bone Metastasis, Yes (%) | 93642 (33.0) | 75821 (33.3) | 17821 (31.7) | <0.001 |
| Brain Metastasis, Yes (%) | 37565 (13.3) | 29214 (12.9) | 8351 (15.0) | <0.001 |
| Lung Metastasis, Yes (%) | 104138 (36.6) | 76277 (33.5) | 27861 (49.2) | <0.001 |
| Liver Metastasis, Yes (%) | 81644 (29.1) | 63075 (28.0) | 18569 (33.5) | <0.001 |
| Primary Cancer Location (%) |  |  |  |  |
| Bladder | 4819 (0.9) | 3791 (0.9) | 1028 (0.9) |  |
| Breast | 34387 (6.5) | 31066 (7.5) | 3321 (3.0) |  |
| CRC | 56763 (10.8) | 48506 (11.6) | 8257 (7.6) |  |
| Gastro-esophageal | 35563 (6.8) | 27269 (6.5) | 8294 (7.6) |  |
| HCC | 7964 (1.5) | 5119 (1.2) | 2845 (2.6) |  |
| Head and Neck | 7647 (1.5) | 6968 (1.7) | 679 (0.6) |  |
| Kidney | 17399 (3.3) | 14803 (3.6) | 2596 (2.4) |  |
| Lung | 210027 (39.9) | 156898 (37.7) | 53129 (48.6) |  |
| Melanoma | 6807 (1.3) | 5755 (1.4) | 1052 (1.0) |  |
| Ovary | 17854 (3.4) | 15027 (3.6) | 2827 (2.6) |  |
| Pancreato-biliary | 59071 (11.2) | 40683 (9.8) | 18388 (16.8) |  |
| Prostate | 28219 (5.4) | 27283 (6.5) | 936 (0.9) |  |

\*Income- omitted 10,615 NA's, Residence-omitted 811 NA's, Bone metastasis -omitted 242413 NA's, Brain metastasis - Omitted 244295 NA's, Lung metastasis - Omitted 245322 NA's, Liver metastasis - Omitted 241850 NA's. HCC: Hepatocellular carcinoma. CRC: Colorectal cancer. NA=Not available

Breast, prostate, and colorectal cancer were less frequent in the early mortality group (3% vs 7.5%, 0.9% vs 6.5% and 7.6% vs 11.6% respectively); in contrast, lung, pancreato-biliary and hepatocellular were more frequent (48.6% vs 37.7%, 16.8% vs 9.8% and 2.6% vs 1.2% respectively). Primary tumor distribution in the whole cohort and in the early mortality group by gender appears in Figure 1.

**Figure 1.**
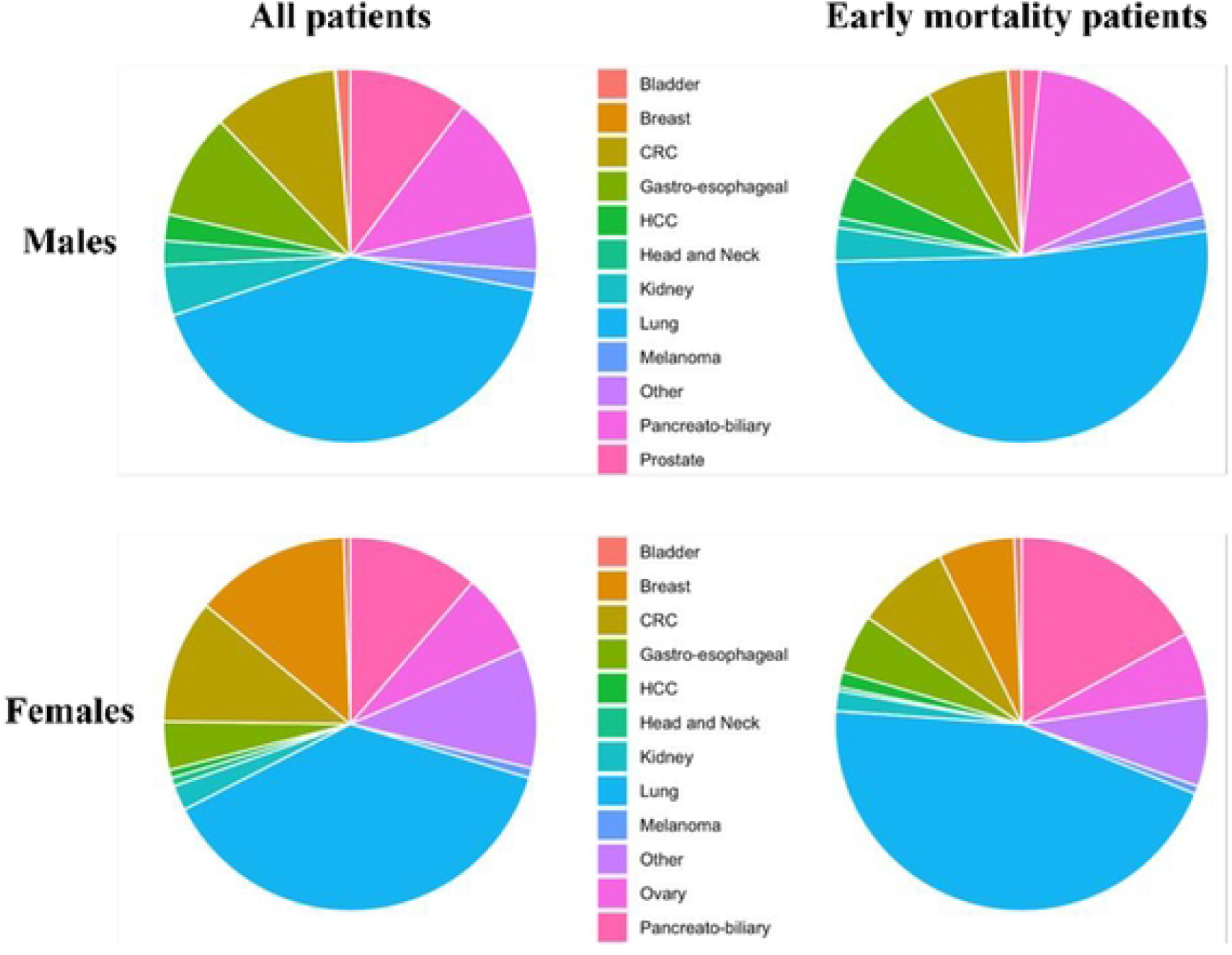
Disease site distribution by gender in the whole cohort and in the early mortality group. Left Panel-525,780 Patients with *de-novo* metastatic cancer were analyzed by disease site origin, separated by gender. Right Panel -109,207 patients which died in the first two months after disease diagnosis analyzed by disease site origin, separated by gender. Primary site color code differs between genders and appears in the middle.

### Causes of death

Within our cohort, 109,207 (20.8%) patients died of cancer within two months of diagnosis. Cancer was the cause of death in 92.1% of early deaths (Figure 2A). The most common non-cancer cause of early death was heart disease n=28,960 (56%) followed by infection n=1,010 (19%). The ten most common non-cancer causes of early death appear in figure 2B.

**Figure 2.**
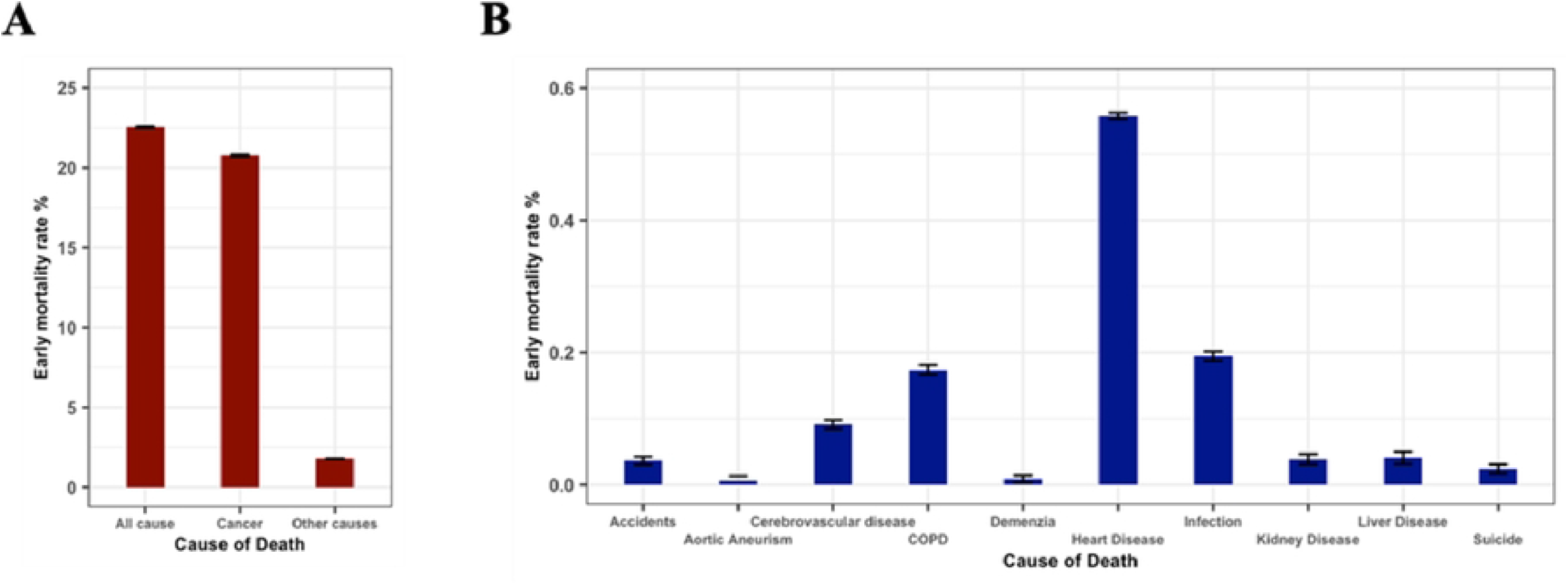
Major causes of early death in de novo diagnosed metastatic cancer patients. All cause and cancer specific early death rates (Figure 2A). Ten major causes of non-cancer early death (Figure 2B). Y axes present percentages in both plots.

### Primary tumor specific early mortality rates

The two-month cancer specific mortality rate was highest in patients with hepatocellular (36%), pancreato-biliary (31%), lung (25%), gastro-esophageal (23%) and bladder (21%) primaries and lowest in patients with prostate (3%), head and neck (9%) and breast (10%) primaries (Figure 3). Black patients had a significantly higher early mortality rate in breast, gastroesophageal, pancreato-biliary, ovarian and kidney cancer, whereas white patients had a significantly higher mortality rate in lung, prostate cancer, and melanoma. These differences remain statistically significant in multivariate analysis. Figure 3 displays early mortality rates by primary tumor, race, and year of diagnosis.

**Figure 3.**
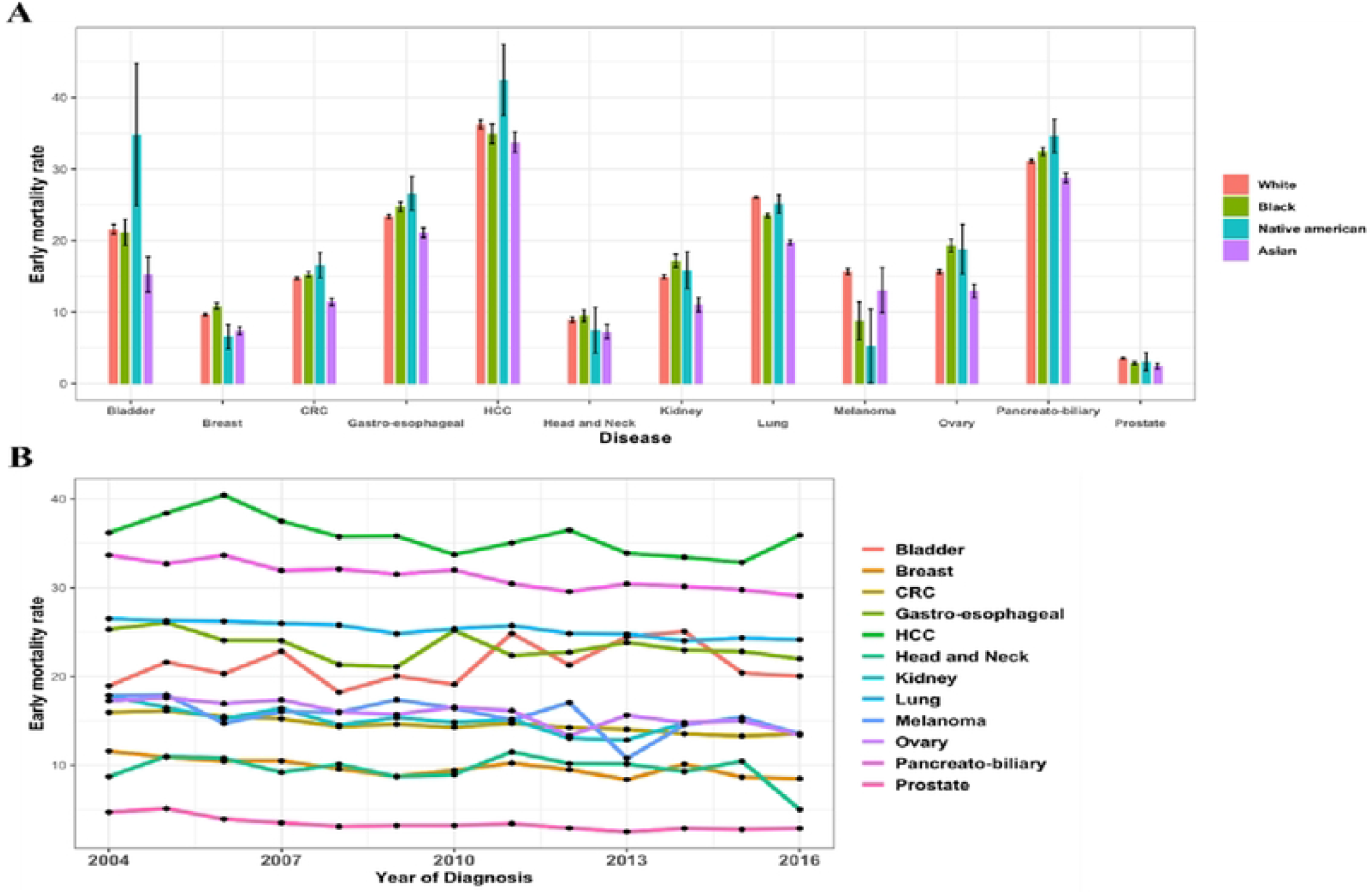
early mortality rates by primary tumor, race, and year of diagnosis. Early mortality rates were calculated separately for each primary tumor group in each race group (Figure 3A). Early mortality trends by primary tumor site and year of diagnosis (Figure 3B). Y axes present percentages.

In most cancer types a statistically significant reduction in early mortality was observed between 2004 and 2016, with the overall two-month cancer specific mortality rate improving during the period of our study from 22.4% in 2004 to 18.8% in 2016. The strongest relative reductions in early mortality were documented in prostate, kidney, and breast cancers.

Cancers without a statistically significant reduction in early mortality include bladder cancer, head and neck cancer and melanoma (Figure 3B).

### Multivariate analysis of all cancers

On multivariate analysis, factors significantly associated with increased two-month mortality (table 2) included site of metastases, particularly liver metastases (data starting from 2010, OR 1.91 CI 1.87-1.96, p<0.001), male sex (OR 1.17 CI 1.16-1.19, p<0.001] income below $65K a year (OR 1.12 CI 1.11.13, p<0.001), race (black and native American) (OR 1.07 CI 1.04-1.09, p<0.001 and OR 1.13 CI 1.02-1.24, p<0.001 respectively), and increasing age (OR 1.03 per year p<0.001).

**Table 2:**
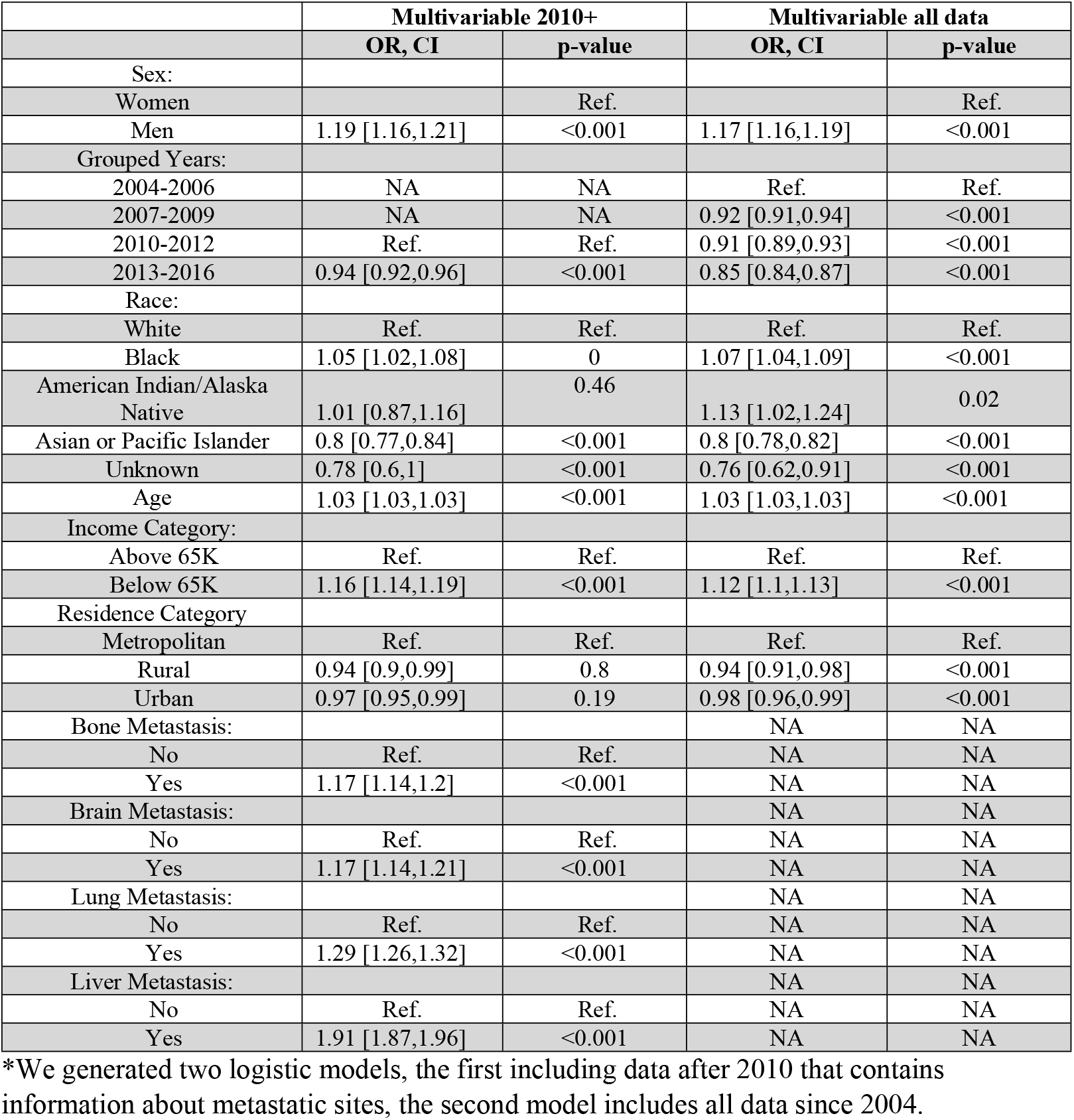
Multivariable logistic regression analysis of the correlates of early mortality in all patients

## Discussion

Our study reports that approximately 20% of patients with biopsy proven *de-novo* metastatic cancer die within two months of diagnosis.

Previous studies of individual cancers have also reported high early mortality rates. An analysis of the national cancer database (NCDB) showed that between 2006 and 2014, 13% of patients with metastatic non-small cell lung cancer (NSCLC) died within 30 days of diagnosis (5). A SEER analysis of patients with metastatic breast cancer showed that in 2013, 13.4% of patients died within one month, while a SEER analysis of patients with pancreatic adenocarcinoma showed that approximately half of patients died within two months of diagnosis (6),(7). These studies demonstrated that early mortality is a prevalent phenomenon, associated with increased age, insurance status, geographic location, and comorbidity burden. Nevertheless, to our knowledge, the scope of early mortality across most solid tumors and the total magnitude of this phenomenon, has never been reported before.

Death so quickly after a cancer diagnosis may be due to causes related to the cancer itself or as result of non-cancer related comorbidities. Interestingly, in our study over 90% of patients that died within two months died from the cancer itself and not from comorbidities. This number is far greater than previously reported (8), likely because our cohort included only patients with metastatic disease who had a histologically confirmed diagnosis. Despite deteriorating rapidly after diagnosis all our patients were deemed well enough to undergo invasive testing during the diagnostic process. We chose to examine two-month mortality because this is the period often required to see an improvement from many cancer therapies and death within this period suggests that patients are unlikely to benefit from most systemic treatments. In our study patients with pancreato-biliary, lung, and liver primaries had the highest proportion of early mortality. Patients with liver metastases were almost twice as likely to suffer early mortality independent of other factors. If properly identified a more personalized diagnostic pathway may improve the care of this large subset of patients with de novo metastatic cancer.

The cancer diagnostic process itself is costly and time consuming. After a patient presents with suspected cancer to a healthcare provider, the health system interval can take a median of 35 days to reach a diagnosis, without taking into consideration another 23 days for molecular testing results required in certain tumor types such as non-small cell lung cancer (9)(10). With contemporary systemic anti-cancer treatments, for many of these patients, early death from cancer is unavoidable even if treatment is initiated immediately after patient presentation. In fact a “waiting time paradox” in which patients with severe symptoms and worse prognosis are more likely to be diagnosed and treated promptly has been described (11) (12) (13). Identifying these patients at high risk for early death may help avoid costly and time-consuming evaluations particularly invasive diagnostic procedures. Early palliative care should be considered even without a tissue diagnosis. On the other hand, it is important to recognize and provide a unique diagnostic approach to the subset of patients that if treated rapidly can achieve a significant and long-term response. For example, in our cohort the absolute number of early deaths is predominantly attributable to lung cancer as this is both the most common *de-novo* metastatic cancer and is also associated with a two-month mortality rate of 25%. Many of these patients succumb rapidly to biologically aggressive disease. Remarkably, many of the advances in molecular targeted therapies and immunotherapies have occurred in lung cancer, including rapid and high response rates (sometimes exceeding 70%) to some molecular targeted therapies or immunotherapies (14). Treatable genomic alterations, such as EGFR mutations, ALK/ROS1 rearrangements and BRAF mutations, can be found in ∼20-30% of metastatic non-small cell lung cancers, suggesting a meaningful percentage of these patients may survive for years if identified timely (14). However, in order for patients who are extremely ill at diagnosis to benefit from targeted therapy, improvements must be made in the clinical uptake and turnaround time of diagnostic and molecular testing (15). Amongst rapidly deteriorating patients, newer and minimally invasive diagnostic tools, such as liquid biopsies, where genomic alterations can be assessed rapidly (16) (17), may identify patients that have a high chance of response to treatments while potentially avoiding more invasive and time-consuming diagnostic interventions such as tissue biopsies. As the price of these tests are constantly decreasing, cost effectiveness will need to be determined but liquid biopsies may identify deteriorating patients who will and equally important, will not, benefit from oncological treatment. For patients who are identified as being at high risk for early death but have a clinical and radiological picture suggestive of metastatic disease from a tumor primary with a high likelihood of response to standard treatment such as breast, prostate, germ cell, or small cell lung cancer standard diagnostic assessments should be fast tracked. For patients at high risk for early mortality with imaging suggestive of metastatic disease from a tumor primary less likely to benefit from standard treatment liquid biopsies should be considered instead of all other diagnostic interventions. These patients don’t stand to benefit from standard non targeted treatment and therefore the identification a targetable mutation quickly is likely their only chance to experience significant life extending treatment. The identification of the small subset of patients with targetable mutations may have far more clinical significance than completing the routine diagnostic process including invasive biopsies for all these patients. For most patients who won’t have actionable mutations it is unlikely that any additional evaluation will affect their prognosis, and they should be referred for early palliative care. As many of these clinical decisions require expertise in clinical oncology, we suggest that a medical oncology consultation should be recommended in patients with suspected metastatic cancer who are at high risk for early mortality at presentation.

Our study has several limitations. We do not have information on important confounders including performance status, comorbidities, socioeconomic environment, and healthcare facilities treated. In addition, we do not have data on physician referral or treatment patterns. Our cohort includes only patients with de novo metastatic cancer and is not applicable to patients with metastatic recurrences.

In conclusion, our study highlights the magnitude of early mortality among *de-novo* metastatic cancer patients and identifies risk factors associated with early mortality including cancer primary, liver and brain metastases, advanced age, and lower income. We showed that most of these patients died of their metastatic disease and not co morbidities. Our study adds to previously described data on early death from individual metastatic solid tumors. Further studies are required to better identify patients at high risk for early mortality, facilitating educated discussion with patients and caregivers. If properly identified a unique and more efficient diagnostic pathway for this patient population may help improve the outcomes of these patients.

## Data Availability

All relevant data are within the manuscript and its Supporting Information files

